# A comparative analysis of statistical methods to estimate the reproduction number in emerging epidemics with implications for the current COVID-19 pandemic

**DOI:** 10.1101/2020.05.13.20101121

**Authors:** Megan O’Driscoll, Carole Harry, Christl A. Donnelly, Anne Cori, Ilaria Dorigatti

**Affiliations:** MRC Centre for Global Infectious Disease Analysis, Department of Infectious Disease Epidemiology,School of Public Health, Imperial College London, London, United Kingdom; Mines ParisTech, Paris 75006 & Université Paris-Saclay, Orsay 91400, France; Department of Statistics, University of Oxford, Oxford,United Kingdom

**Author notes:** Equal contribution (Megan O’Driscoll, Carole Harry). Correspondence &.

**Keywords:** Outbreak analysis;, SARS-CoV-2, reproduction number, estimation method comparison, emerging epidemics

## Abstract

As the SARS-CoV-2 pandemic continues its rapid global spread, quantification of local transmission patterns has been, and will continue to be, critical for guiding pandemic response. Understanding the accuracy and limitations of statistical methods to estimate the reproduction number, R_0_, in the context of emerging epidemics is therefore vital to ensure appropriate interpretation of results and the subsequent implications for control efforts. Using simulated epidemic data we assess the performance of 6 commonly-used statistical methods to estimate R_0_ as they would be applied in a real-time outbreak analysis scenario – fitting to an increasing number of data points over time and with varying levels of random noise in the data. Method comparison was also conducted on empirical outbreak data, using Zika surveillance data from the 2015–2016 epidemic in Latin America and the Caribbean. We find that all methods considered here frequently over-estimate R0 in the early stages of epidemic growth on simulated data, the magnitude of which decreases when fitted to an increasing number of time points. This trend of decreasing bias over time can easily lead to incorrect conclusions about the course of the epidemic or the need for control efforts. We show that true changes in pathogen transmissibility can be difficult to disentangle from changes in methodological accuracy and precision, particularly for data with significant over-dispersion. As localised epidemics of SARS-CoV-2 take hold around the globe, awareness of this trend will be important for appropriately cautious interpretation of results and subsequent guidance for control efforts.

**Significance Statement:** In line with a real-time outbreak analysis we use simulated epidemic data to assess the performance of 6 commonly-used statistical methods to estimate the reproduction number, R_0_, at different time points during the epidemic growth phase. We find that estimates of R_0_ are frequently overestimated by these methods in the early stages of epidemic growth, with decreasing bias when fitting to an increasing number of time points. Reductions in R_0_ estimates obtained at sequential time points during early epidemic growth may reflect increased methodological accuracy rather than reductions in pathogen transmissibility or effectiveness of interventions. As SARS-CoV-2 continues its geographic spread, awareness of this bias will be important for appropriate interpretation of results and subsequent guidance for control efforts.

## Introduction

The reproduction number, R, is a key epidemiological parameter that quantifies the average number of new infections caused by a single infected individual. When a pathogen emerges in an entirely susceptible population this parameter is referred to as the basic reproduction number, R_0_. When some population-level immunity exists, the parameter is referred to as the effective reproduction number, R_e_. These reproduction numbers provide valuable information about a pathogen’s potential for spread in a population and the associated implications for control efforts (1–3). Reproduction number estimates for any one pathogen are time and context specific, with factors such as contact patterns, population immunity, and behavioral change contributing to variability in these estimates (4, 5). For these reasons, the reproduction number is usually monitored over time to track the progress of an outbreak. The instantaneous reproduction number, R_t_, estimates the average number of secondary infections generated by an infected individual at time t and sequential estimates over the course of an outbreak can provide valuable insights into the need for interventions and/or the effectiveness of control programmes already in place.

Numerous mathematical and statistical methods have been developed to estimate the reproduction number of an emerging pathogen (6–12). The choice of method to be used largely depends on the available data, with more data typically allowing the parameterization and use of more complex methods. However, in the early stages of an outbreak, epidemiological data are often sparse and highly uncertain. In the case of emerging pathogens, data can be particularly limited, as surveillance systems may be unprepared for the detection and reporting of a new pathogen. In addition, limited understanding of the dynamics of a new pathogen can often inhibit the parameterization and use of more complex transmission models. This has particularly been the case for SARS-CoV-2, which was first reported in China in December 2019 and has since reached more than 200 countries and territories, as of May 2020 (13). Though new evidence is rapidly emerging on aspects of transmission dynamics such as generation intervals and the proportion of asymptomatic infections (14), many uncertainties remain, including the duration of immunity, the role of seasonality on transmission and the effects of immunological cross-reactivity with endemic human coronaviruses (HCoV). Incorporating such uncertainties into mechanistic models of transmission can prove challenging in early outbreak contexts, where statistical methods are often the only available tool to infer the level of transmission from a limited amount of data, such as a time series of reported case numbers. Quantifying local transmission patterns of SARS-CoV-2 has been, and will continue to be, critical for guiding the pandemic response (15–18). Understanding the accuracy and limitations of these methods in the context of real-time outbreak analysis scenarios is crucial in order to optimally inform outbreak response activities, including the need for new control efforts or the relaxation of efforts already in place.

Here, we conduct a comparative performance analysis of 6 commonly-used statistical methods to estimate the reproduction number at sequential time points during the early stages of an emerging epidemic, in line with a real-time outbreak analysis scenario. We assess the performance of methods at each time point on simulated epidemic data with varying levels of random noise. We then apply each method to case-notification data provided by national surveillance systems in Latin America and the Caribbean during the 2015–2016 Zika epidemic, providing new insights into the comparability of estimates obtained with different methods.

## Results

Biases in estimates of R_0_ (estimated R_0_ – actual R_0_) when fitted to increasing time points in the case time series (6, 9, 12 and 15 weeks) of simulated data are shown in Figure 1 and Supplementary Figure S2 for the scenario of data with no added noise, i.e. assuming that a constant proportion of infected individuals are detected and reported accurately in the case time series. Estimates of R0 were frequently over-estimated at all time points assessed, the magnitude of which was generally greater for higher values of actual R_0_ (Figure S2). As expected, higher values of R_0_ used for data simulation generally resulted in epidemics that peaked at earlier time points, subsequently resulting in a reduction in the number of epidemic simulations where the growth phase continues beyond 9, 12, 15, etc., weeks, and an under-representation of high R_0_ simulations at the later epidemic stages assessed (Figure S2). To ensure a systematic analysis of methodological performance at sequential time points in the epidemic time series, assessment of performance was restricted to epidemic simulations that peaked ≥15 weeks, highlighted by the blue points in Figure S2.

**Figure 1.**
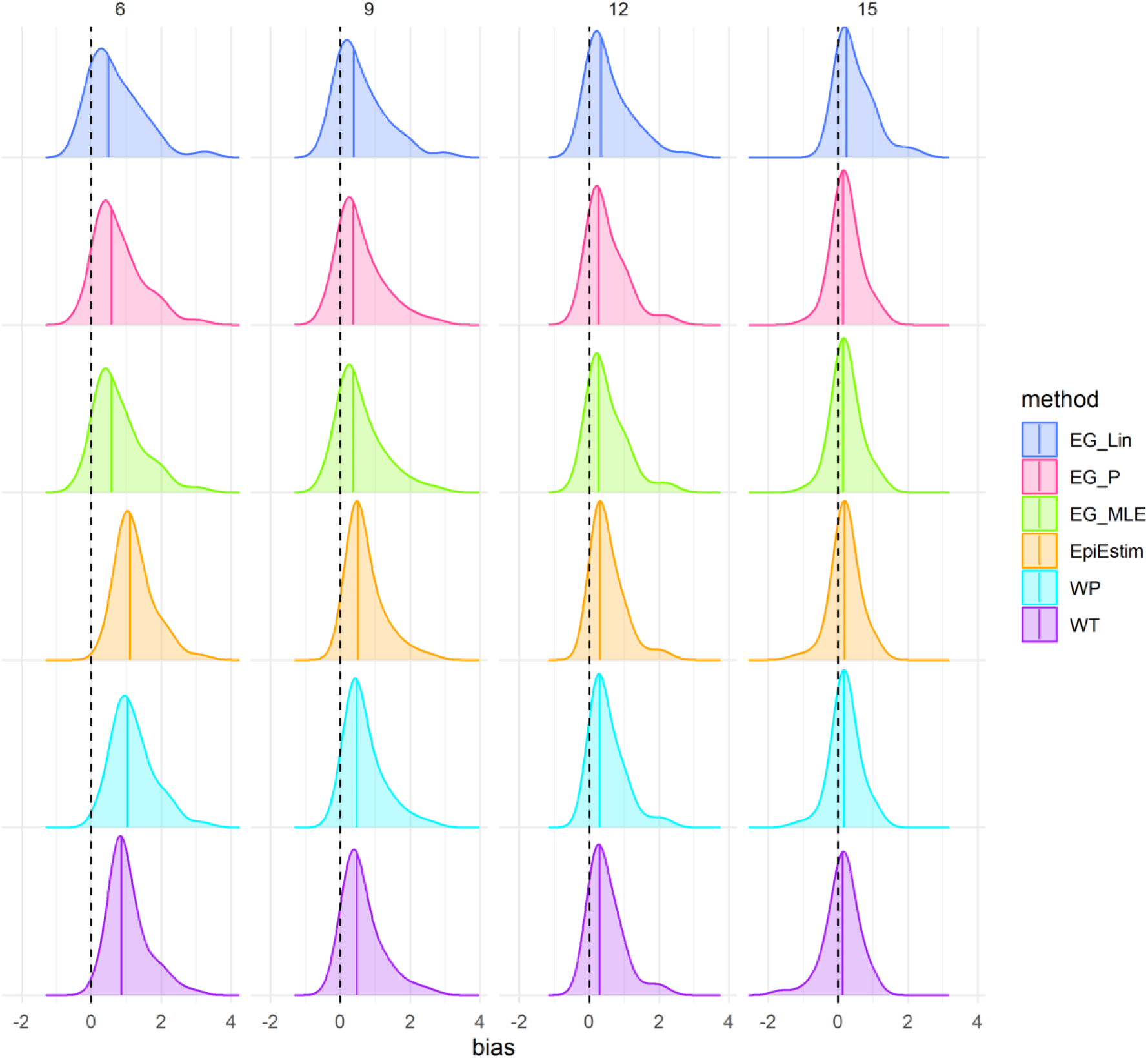
Density distributions of bias in R_0_ estimates (estimated R0 – actual R_0_) obtained when fitting to the case time series on simulated data, without noise, by method and time point (in weeks), using only results from simulations that peaked ≥15 weeks (N = 146). Columns represent the number of datapoints (weeks) each method was fitted to in the case time series (6,9,12,15 weeks, approximating to 2,3,4,5 generation times), and colours represent the method. Black dashed lines highlight the ideal bias value of zero and coloured lines represent method-specific values of median bias. Method abbreviations: Linear exponential growth rate method (EG_Lin); Poisson exponential growth rate method (EG_P); maximum likelihood exponential growth rate method (EG_MLE); White and Pagano method (WP), Wallinga and Teunis (WT).

Average bias between estimated and actual R0 values was highest in the earliest stages assessed for all methods considered (Figure 1 and S3), with mean absolute differences between estimated and actual R0 values ranging from 0.75 (linear exponential growth) to 1.25 (EpiEstim) when fitting to the first 6 weeks of data (approximately 2 disease generations) of the epidemic growth phase with no added noise. Bias decreased substantially when fitting to an increasing number of time points and ranged from 0.33 (Poisson exponential growth, maximum-likelihood exponential growth and EpiEstim) to 0.51 (linear exponential growth) at 15 weeks (approximately 5 disease generations) in the scenario of no random noise. Density distributions of bias from the scenarios of simulated epidemic data with added noise are shown in Figure S6. For epidemic simulations with Poisson noise, mean absolute bias ranged from 0.88 (linear exponential growth) to 1.64 (Wallinga and Teunis) at 6 weeks and from 0.36 (Poisson and maximum-likelihood exponential growth) to 0.49 (linear exponential growth) at 15 weeks. Similarly, for epidemic simulations with negative binomial noise, mean absolute bias ranged from 1.07 (linear exponential growth) to 2.60 (Wallinga and Teunis) at 6 weeks and from 0.48 (Poisson exponential growth) to 0.80 (Wallinga and Teunis) at 15 weeks. The relationship between estimated R_0_ and actual R_0_ for individual simulations by method and time point are shown in Figures S3–S5 for the scenarios of data with no added noise, Poisson noise and negative binomial noise, respectively.

Coverage of actual R_0_ values, i.e. the proportion of simulations in which the 95% confidence interval contained the actual value of R_0_, decreased when fitted to an increasing number of time points in the case time series for all methods assessed and in all noise scenarios (Figure 2). This decrease in coverage of actual R0 values corresponds to reduced uncertainty, i.e. width of the 95% confidence intervals, surrounding the estimates of all methods when fitted to an increasing number of time points, as shown in Figure 2. In the scenario of data with no noise, coverage ranged from 73% (linear exponential growth and EpiEstim) to 97% (Poisson and maximum-likelihood exponential growth) at 6 weeks and from 23% (linear exponential growth) to 73% (Wallinga and Teunis) at 15 weeks. When fitted to simulations with Poisson random noise, coverage ranged from 57% (EpiEstim) to 94% (Wallinga and Teunis) at 6 weeks and from 49% (EpiEstim) to 66% (Wallinga and Teunis) at 15 weeks. For simulations with negative binomial random noise, coverage ranged from 52% (EpiEstim) to 99% (linear exponential growth) at 6 weeks and from 41% (Poisson and maximum likelihood exponential growth) to 77% (linear exponential growth) at 15 weeks. A significant increase in the levels of uncertainty in the presence of increasing random noise was observed for each of the exponential growth methods (Figure 2). This increase in uncertainty was most notable for the linear exponential growth method, with average confidence interval widths (averaged across all time points assessed) of 0.79, 2.60 and 4.48 when fitted to data with no noise, mild noise (Poisson) and high noise (negative binomial), respectively. Minor increases in the level of uncertainty were observed for the White and Pagano (2.26, 2.40, 2.52) and EpiEstim (1.56, 1.65, 1.72) methods when fitted to data with increasing random noise, whilst no significant differences in levels of uncertainty were observed for the Wallinga and Teunis method (3.75, 3.64, 3.40) with the addition of random noise to the data.

**Figure 2.**
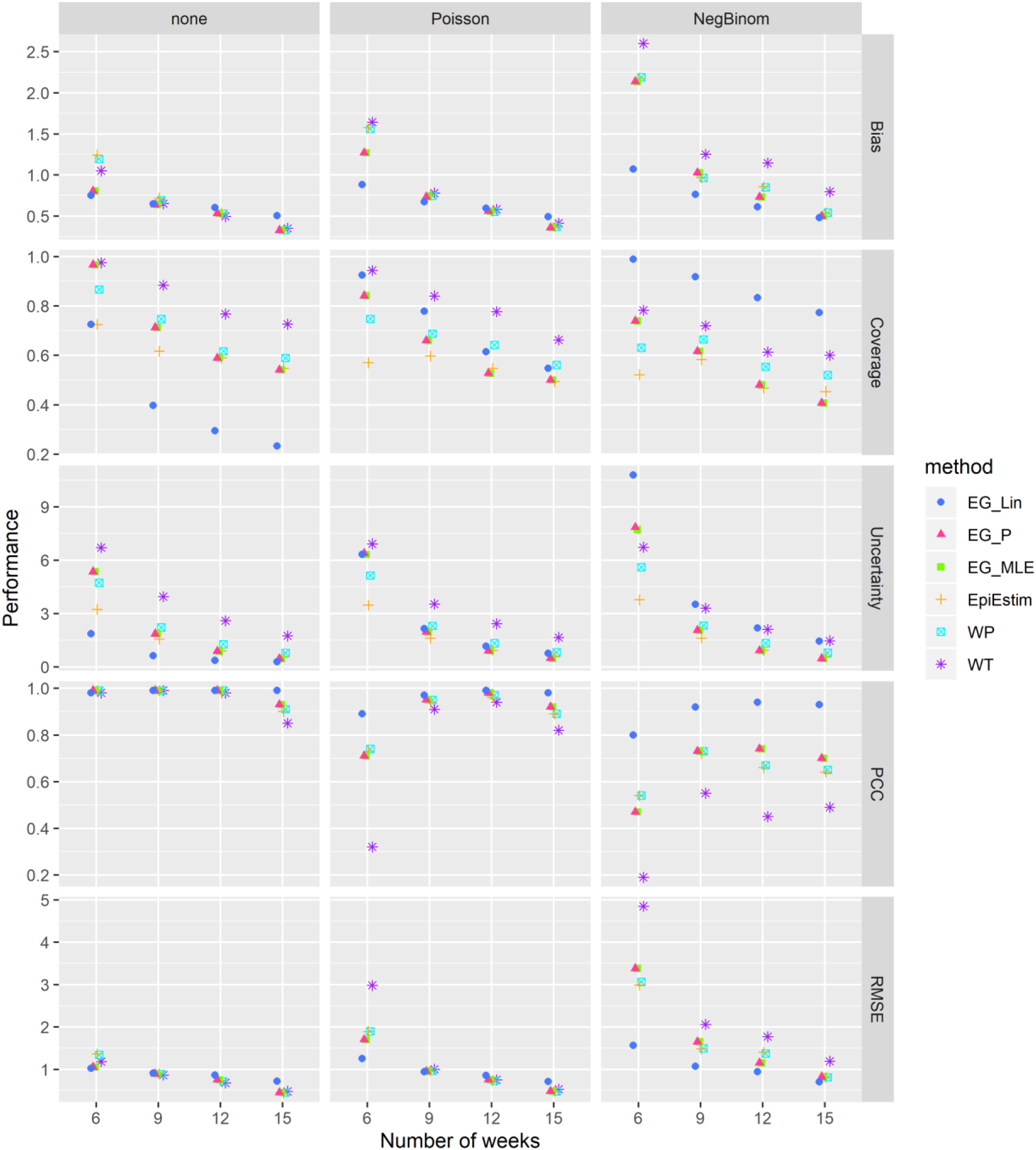
Comparative analysis of the performance of the 6 methods at different stages of the epidemic. Columns represent the three data noise scenarios explored (no noise, Poisson noise and negative binomial noise). Rows represent different performance metrics: Absolute bias (the absolute average difference between estimated and true R_0_ values), Uncertainty (95% confidence interval width), Coverage (proportion of times in which the true R_0_ value is within the estimated 95% confidence intervals), Pearson correlation coefficient (PCC), and root mean squared error (RMSE). Method abbreviations: Linear exponential growth rate method (EG_Lin); Poisson exponential growth rate method (EG_P); maximum likelihood exponential growth rate method (EG_MLE); White and Pagano method (WP), Wallinga and Teunis (WT).

The PCC between estimated and actual R_0_ values was close to 1 for all methods in the scenario of data with no added noise at all time points, though a slight reduction in this correlation was observed at 15 weeks for all methods except linear exponential growth (Figure 2). Reduced correlation between estimated and actual R_0_ was observed for the scenarios of data with random noise at all stages of the epidemic, the most significant reduction of which was observed at the earliest time point assessed (6 weeks). The RMSE decreased when fitted to increasing numbers of data points for all methods considered and in all data noise scenarios. Trends in method-specific RMSE were similar for data with no noise and Poisson noise, though a slight increase in RMSE was observed at 6 weeks for all methods in the Poisson noise data scenario compared to data with no noise. RMSEs increased significantly in the presence of negative binomial noise for all methods, particularly in the earlier epidemic stages assessed.

Figure 3 shows the R_0_ estimates obtained from fitting each of the 6 methods to empirical outbreak data, with the examples of French Guyana, Martinique, Puerto Rico and the US Virgin Islands highlighting empirical outbreaks where estimates of R_0_ decreased when fitting to an increasing number of time points in the case time series during the phase of early epidemic growth. The estimates obtained for all Latin American and the Caribbean countries are shown in Figure S7. R_0_ estimates were generally higher in the early stages of the epidemic for all methods assessed, with estimates decreasing gradually over time. Confidence interval widths were also observed to generally decline when fitted to an increasing number of datapoints. Frequent inconsistencies between estimates of R_0_ produced by different methods on the same data were observed, particularly during the early growth phase of the epidemics, e.g. French Guyana, Martinique and Brazil.

**Figure 3.**
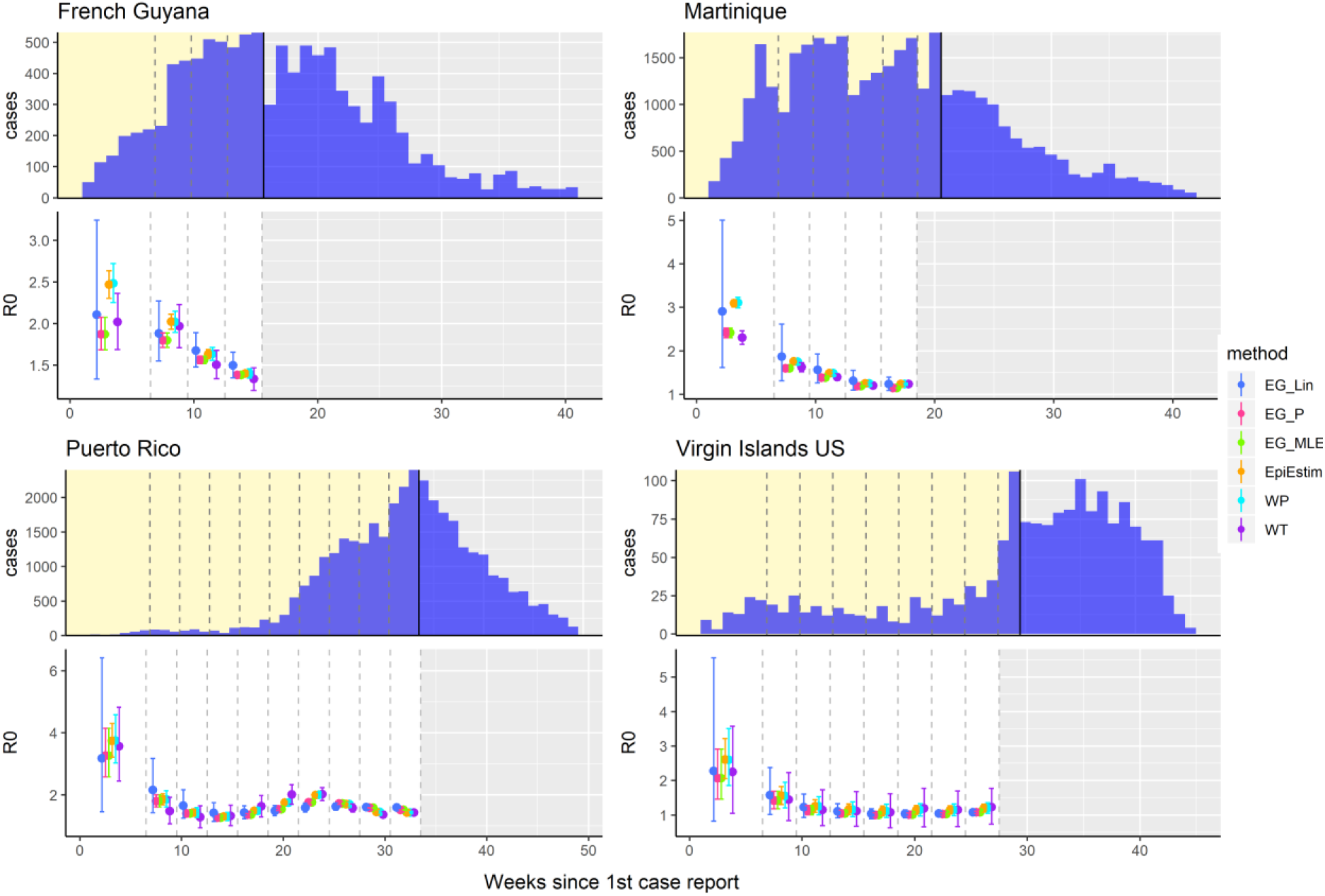
R_0_ estimates obtained from each of the six methods fitted at different stages of the 2015–2016 Zika epidemics in French Guyana, Martinique, Puerto Rico, and the US Virgin Islands. The top panel for each country shows the time series of reported Zika cases, with dashed lines showing the different stages at which each method was fitted to the data(first 6, 9, 12, etc., weeks) up to the peak of the epidemic, marked by the black line. The bottom panel for each country shows the mean and 95% confidence intervals of the R_0_ estimates produced with each method fitted to each time series. Method abbreviations: Linear exponential growth rate method (EG_Lin); Poisson exponential growth rate method (EG_P); maximum likelihood exponential growth rate method (EG_MLE); White and Pagano method (WP), Wallinga and Teunis (WT).

Density distributions of bias in R_0_ estimates when fitting to epidemic simulations using different generation time distributions, are shown in Figure 4. Trends in bias over time were similar when fitted to simulations for Zika and Ebola generation time distributions. Larger average bias was observed for all methods and time points when fitted to simulations for the SARS generation time distribution, which also decreased when fitted to an increasing number of data points in the case time series. Significantly higher average bias was observed for R_0_ estimates produced by the White and Pagano and Wallinga and Teunis methods, relative to other methods for the SARS simulations at all time points assessed.

**Figure 4.**
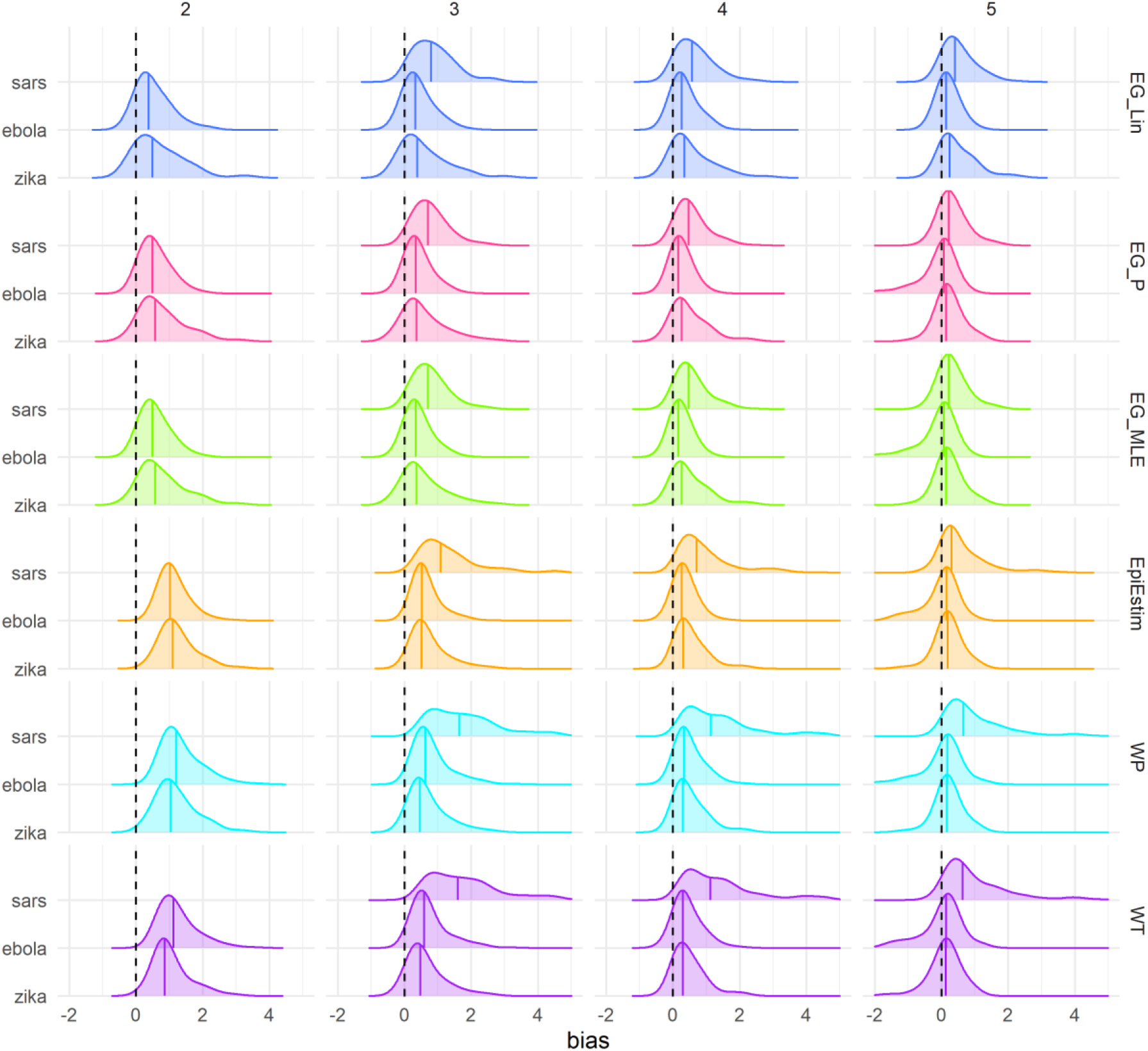
Density distribution of bias in R_0_ estimates (estimated R_0_ – actual R_0_) obtained when fitting to the case time series of simulated data, without noise, by method and time point (in approximate generations) using only results from simulations that peaked ≥15 weeks (N = 146). Columns represent the approximate number of disease generations fitted to in the case time series, and colours represent the method. Black dashed lines highlight the ideal bias value of zero and coloured lines represent method-specific values of median bias. The generation time distribution used, both for data simulation and method fitting, is shown on the y-axis. Mean and standard deviation in days for generation time distributions used: Zika (20±7.4); Ebola (16±9.3); SARS (8±3.8). Method abbreviations: Linear exponential growth rate method (EG_Lin); Poisson exponential growth rate method (EG_P); maximum likelihood exponential growth rate method (EG_MLE); White and Pagano method (WP), Wallinga and Teunis (WT).

## Discussion

In the early stages of an outbreak response, efforts are often dedicated to estimating pathogen transmissibility in order to provide information on the potential for spread in the current population and to inform the type and scale of interventions required for control (19, 20). Our results show variable accuracy of R_0_ estimates obtained, both between individual methods as well as across different stages of the early growth phase of the epidemic. Estimates of R_0_ obtained for the earliest stages of the epidemic assessed in this analysis (i.e. at 6 and 9 weeks, corresponding to approximately 2 and 3 generation time intervals respectively) were associated with larger bias and uncertainty for all methods assessed. Using simulated epidemic data, we found that estimates of R_0_ frequently overestimated the actual R0 value used in the simulation process, particularly in the early fitting stages, 6 and 9 weeks, even when the true case time series (i.e. a constant proportion of infected individuals that is accurately detected and reported) is observed. Estimates of R_0_ became increasingly accurate for all methods when fitted to an increasing number of weeks in the case time series, with the average absolute bias ranging from 0.33 to 0.51 across methods when fitted to 15 weeks (approximately 5 generations for Zika) of data in the scenario with no noise. Higher average absolute bias was observed for all methods when fitting to simulated data with added random noise, particularly in the earlier stages assessed (Figure S6).

Whilst the over-estimation of R_0_ in the early stages of an epidemic may be preferable to underestimation for purposes relating to the planning of outbreak response activities, reductions in the magnitude of this over-estimation over time can potentially lead to misconceptions about trends in transmissibility. The trend of decreasing average R0 estimated in the early stages of epidemic growth was also observed for some countries when fitting to national case surveillance data from the 2015- 2016 Zika epidemic in Latin America and the Caribbean (e.g. French Guyana, Martinique, Puerto Rico and the US Virgin Islands as shown in Figure 3). The limited efficacy of existing interventions against vector-borne pathogens such as Zika suggest the possibility that methodological bias could explain these trends rather than reductions in transmission at that time. In addition, early analysis of the SARS-CoV-2 epidemic in China estimated a declining trend in the reproduction number from 7.93 (95% CI: 5.00–12.00) on the 29^th^ December 2019 to 2.60 (95% CI: 0.57–5.17) on the 18^th^ January 2020 using the Wallinga and Teunis method, attributing this reduction to the effectiveness of prevention and control measures taken at that time (21). However, the largest decrease in R estimated by Liu et al., was observed between the 29^th^ of December and the 2^nd^ of January 2020, prior to the implementation of significant interventions, from which point the estimates of R largely stabilised for the remainder of the period of estimation (21). Apparent trends of decreasing R_0_ during early epidemic growth can potentially be misinterpreted as larger-than-actual decreases in pathogen transmissibility, which may lead to overestimation of the potential impact of interventions and longer than expected times to epidemic extinction. Awareness of this bias is crucial for estimating transmissibility in the early stages of an epidemic and accurately interpreting the subsequent implications for control (22).

In the current context of the SARS-CoV-2 pandemic, caution is required when using these methods for the estimation of the reproduction number. As countries expand surveillance systems to better manage the pandemic, the assumption of constant reporting of cases implicit in these methods likely do not hold true until the capacity of the surveillance system has stabilised, and a consistent case definition is applied. In these contexts, whilst testing capacity is growing, the use of hospital admission or death data, where available, may be preferable for inferring R_0_ if the reporting of these data is believed to be constant in time. Delays in the time from infection to hospitalisation and/or death, however, result in significant lags between when the transmission events occur and when they can be quantified. Regardless of the metric used for estimation of R_0_, we urge caution when interpreting short-term fluctuations in these estimates as variable data quality and methodological accuracy may play a role in these trends.

Previous work by Obadia et al. compared estimates of the reproduction number produced by 4 generic methods (exponential growth, maximum likelihood, sequential Bayesian and time-dependent methods) (23). However, the comparison was conducted at a single time-point in the epidemic case time series (24 days, the equivalent of 6 generation intervals for a typical influenza generation time distribution) and new methods have since been developed to estimate the reproduction number. We observed varying strengths and limitations associated with each of the methods considered here and the sensitivity of these factors to levels of random noise and the amount of available data. While the linear exponential growth rate method performed poorest in terms of coverage of actual R_0_ values when fit to simulated data with no added noise, it had the highest coverage of actual R_0_ values when fit to data with added random noise, driven by increased confidence interval widths in the presence of overdispersion. Conversely, the Wallinga and Teunis method had the highest coverage of actual R_0_ values when fitted to data with no noise and mild (Poisson) noise but had the largest average bias across all time points when fitted to data with high overdispersion (negative binomial noise).

There are a number of limitations in our analysis, including the simplicity inherent in the assumptions of homogenous mixing in fully susceptible and closed populations that were used to simulate the case data. These assumptions do not reproduce the sub-exponential growth dynamics that can occur in empirical epidemics as driven by complex spatial structures, local contact networks and other socio-behavioural factors not accounted for in this analysis (24, 25). The results of this analysis are based on weekly epidemic case-time series, which may not fully reflect results obtained when daily case reports are available. However, whilst the larger number of datapoints provided by daily time series may potentially improve methodological performance in the early epidemic stages, daily case counts may be subject to increased errors/overdispersion and substantial day-of-the-week effects. In addition, noise was added to the case time series in the form of random errors to reflect stochasticity in the infection and reporting processes, inherently assuming the noise to be random. Systematic forms of noise/bias, such as reporting delays, changes in health-seeking behaviour or case definitions, incorrect generation time assumptions etc., are not accounted for in this analysis. Here, we show that R_0_ estimates frequently overestimate the actual R_0_ values even when the true case time series is observed (i.e. a constant proportion of infections that is accurately detected and reported), when the true generation time is known, and when the assumption of exponential growth is met (both in the simulation model and in the estimation method).

In this analysis we highlight the varying strengths and limitations of 6 commonly-used statistical methods for estimating R_0_ in emerging epidemics. We show how the performance of these methods can vary over time when fit to increasing amounts of case data, in line with a real-time outbreak analysis scenario, and demonstrate the sensitivity of methodological performance to varying levels of random noise in the data. This has important implications for the ongoing SARS-CoV-2 pandemic as we show that true changes in transmissibility in the early stages of epidemic growth may be difficult to disentangle from changes in methodological accuracy and precision, particularly for data with significant overdispersion. Cautious interpretation is warranted when using these methods to infer transmission patterns in the early stages of epidemic growth, as apparent declines in R_0_ estimates may be mis-attributed to the effectiveness of control efforts and lead to incorrect conclusions about the course of the epidemic. Several generations of pathogen transmission may need to be observed for accurate R_0_ estimation by these methods. An awareness of this trend in bias over time is crucial for appropriate interpretation of R_0_ estimates and any subsequent implications for the planning of outbreak response activities.

## Methods

### Data Simulation

Incidence data were simulated using a deterministic compartmental SEIR (susceptible, exposed, infectious, recovered) model of transmission, assuming a time-constant basic reproduction number, R_0_. Values of R_0_ were randomly selected from a truncated normal distribution with a mean of 2, standard deviation of 2, truncated between 1.2 and 8 (Figure S1 in supplementary information). Using the example of Zika, we assumed average incubation and infectious periods of 14 and 6 days, respectively, for data simulation, giving an average generation time of 20 days, standard deviation 7.4, which we assumed to be known for model fitting (Figure S1 in supplementary information) (26). We assumed there was no transmission before the onset of symptoms. We also assumed entirely susceptible closed populations with randomly selected sizes ranging in multiples of 10, from 10^3^ to 10^6^, and assumed homogenous mixing in the population. The epidemic was seeded with a random number of between 1 and 5 initially infected individuals. Daily incidence data were simulated for a period of 2 years which was then aggregated to weekly data for consistency with the empirical data reported during the 2015–2016 Zika epidemic in Latin America and the Caribbean. We assumed that 20% of all infections resulted in reported case-notifications which was constant over time. 250 unmitigated epidemic simulations were generated, each of which was fitted to under 3 different scenarios of random noise – no noise, mild noise and high noise. To represent stochasticity in the reporting and infection processes underlying the observed case incidence, Poisson (mild noise) and negative binomial (high noise, dispersion parameter=3) distributed random errors were separately added to the simulated epidemic.

### Statistical methods

#### 1. Exponential Growth methods

The exponential growth rate, *r*, is the per capita change in the number of new cases per unit of time. This growth rate can be linked to the basic reproduction number, *R_0_*, through the moment generating function of the generation time distribution, *g(a)*, (27) as shown in equation 1:

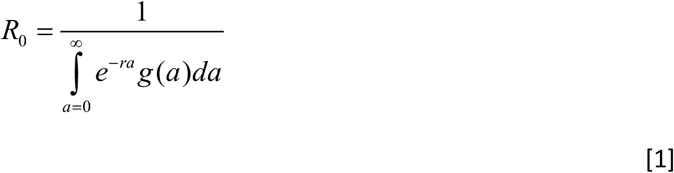

As the name suggests, this method assumes the epidemic to be growing at an exponential rate and with a known generation time distribution, *g(a)*. This method can be applied to the case time series using Poisson regression which we denote as “EG_P”, or on the logarithm of the case time series using linear regression, denoted “EG_Lin”. We apply both Poisson and linear regressions in this analysis. The 95% confidence interval around the central R_0_ estimates were derived by taking the bounds of the 95% confidence interval for the estimate of r, which were transformed to R0 values as in equation 1.

In addition, we applied a maximum likelihood version of the method to estimate both the exponential growth rate *r* and *I_0_*, the initial number of infected individuals, assuming that incidence at time *t*, It, is Poisson distributed with mean *I*_0_^*rt*^, which we denote as “EG_MLE”.

#### 2. White and Pagano maximum likelihood method

The White and Pagano method uses a maximum likelihood framework with the log-likelihood (LL) shown in equation 2. The mean case incidence, *µt*, is defined as shown in equation 3, where *It* denotes the observed incidence at time *t* and *g(s)* denotes the generation interval distribution.

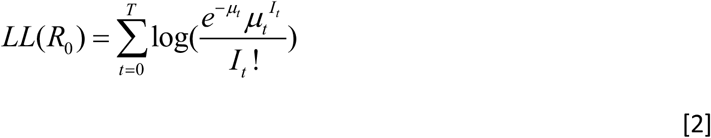

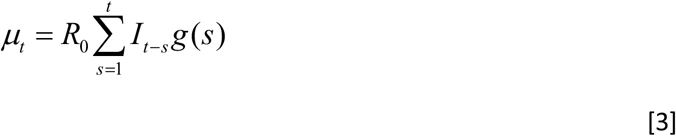

This method assumes an infinite number of susceptible individuals, with no importation or exportation of cases, and that the number of secondary cases generated by an index case is Poisson distributed (11).

#### 3. EpiEstim maximum likelihood method

EpiEstim was originally designed to estimate time-varying reproduction numbers, Rt, but can also be applied to estimate what can be interpreted as the basic reproduction number, R_0_ (12). As in the White and Pagano method, this method uses maximum likelihood estimation assuming that case incidence is Poisson distributed with mean *µt*, as shown in Equations 2 and 3. The posterior distribution of R_0_ is calculated in a Bayesian framework with a gamma distributed prior distribution for R_0_ (12). Here the generation time distribution is used as an approximation for the infectivity profile of cases, i.e. the probability of an infected individual generating another case. This method assumes no importation or exportation of cases. A recent update to this method has been developed to account for imported cases (28), however, we restrict our current analysis to that of closed systems.

#### 4. Wallinga and Teunis maximum likelihood method

The Wallinga and Teunis method is also a time-dependent method which computes the reproduction number by averaging over all networks of transmission that are compatible with observed data (10). Here, the relative likelihood that case *i* has been infected by case *j*, normalized by the likelihood that case *i* has been infected by any other case *k* is:

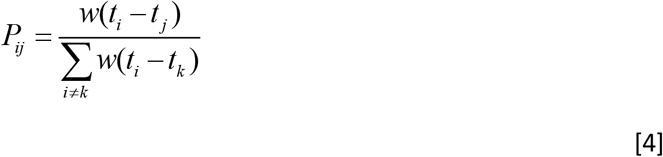

where *w* is the generation time distribution and *t_i_*-*t_j_* is the difference in time of symptom onset between cases *i* and *j*. Though the Wallinga and Teunis method is most often used to estimate the case reproduction number, which accounts for past, present and future cases, in this analysis we apply the method only to cases observed at the current time point in order to assess real-time methodological performance. Average R^0^ estimates were obtained by smoothing the time-dependent estimates across the period of interest, which were corrected to account for secondary cases that are not yet observed (9).

#### Assessing performance on simulated data

Estimating an average, fixed R_0_, we assessed the performance of each statistical method by fitting to the simulated epidemic data at sequential time points in the epidemic growth phase, in line with a real-time outbreak analysis scenario. A fixed gamma distribution for the generation time interval with mean of 20 days and standard deviation of 7.4 days was assumed to be known for all methods (26). We initiated the analysis 6 weeks into each epidemic, fitting to the first 6,9,12,15, etc., weeks (approximating to 2,3,4,5, etc., generation times), up to the peak of the epidemic, simply defined as the week with the maximum number of cases from the entire epidemic curve. The performance of each method was assessed at each of these stages, with methods being fit to an increasing number of data points. We used a number of metrics to assess method performance: bias, calculated as the difference between the estimated and true R_0_ values; coverage, calculated as the proportion of times the estimated 95% confidence intervals included the true R_0_ value; Pearson correlation coefficient (PCC); width of 95% confidence intervals; and the root mean square error (RMSE). To ensure a systematic comparison of performance we only compared the results from sections of the epidemic time series where all methods were able to produce estimates of R_0_.

A sensitivity analysis was conducted to assess how methodological performance might differ for pathogens with different generation time distributions to that of Zika. In the same manner as for the Zika epidemic simulations, 250 unmitigated epidemic simulations were generated using an ‘Ebola-like’ generation time distribution with a mean of 16 days and standard deviation of 9.3 days (assuming incubation and infectious periods of 11 and 5 days) and a ‘SARS-like’ generation time distribution with mean of 8 days and standard deviation 3.8 days (assuming incubation and infectious periods of 5 and 3 days). In line with weekly case reporting that occurred during the Zika epidemic, the mean generation time was approximated to the nearest number of weeks (i.e. ∼3 weeks for Zika, ∼2 weeks for Ebola and ∼1 week for SARS) and fitted to case data approximating to the first 2,3,4 and 5 generations times. For the ‘Ebola-like’ generation time, this was approximated to the first 4,6,8 and 10 weeks of data and to the first 2,3,4 and 5 weeks for ‘SARS-like’ generation time.

#### Fitting to empirical data from the 2015–2016 Zika epidemic in Latin America and the Caribbean

We applied each method to weekly case-notification data from the 2015–2016 Zika epidemic in Latin America and the Caribbean. The data were available at the national level for 37 countries, 3 of which reported their peak incidence within 6 weeks of the first reported Zika case and were subsequently removed from the analysis. In the same manner as the simulated data, we applied each of the 6 methods to an increasing number of data points (using the first 6,9,12,15, etc., weeks), up to the peak of the epidemic.

## Data Availability

All data and code necessary to reproduce this analysis are available at https://github.com/meganodris/R0-methods-comparison

## Acknowledgments

We thank Dr Zulma Cucunubá for sharing an updated version of the Zika incidence data from Latin America and the Caribbean (26).

## Data Statement

The data and R code necessary to reproduce this analysis are available on GitHub: https://github.com/meganodris/R0-methods-comparison

## Supplementary Information

**Figure S1.**
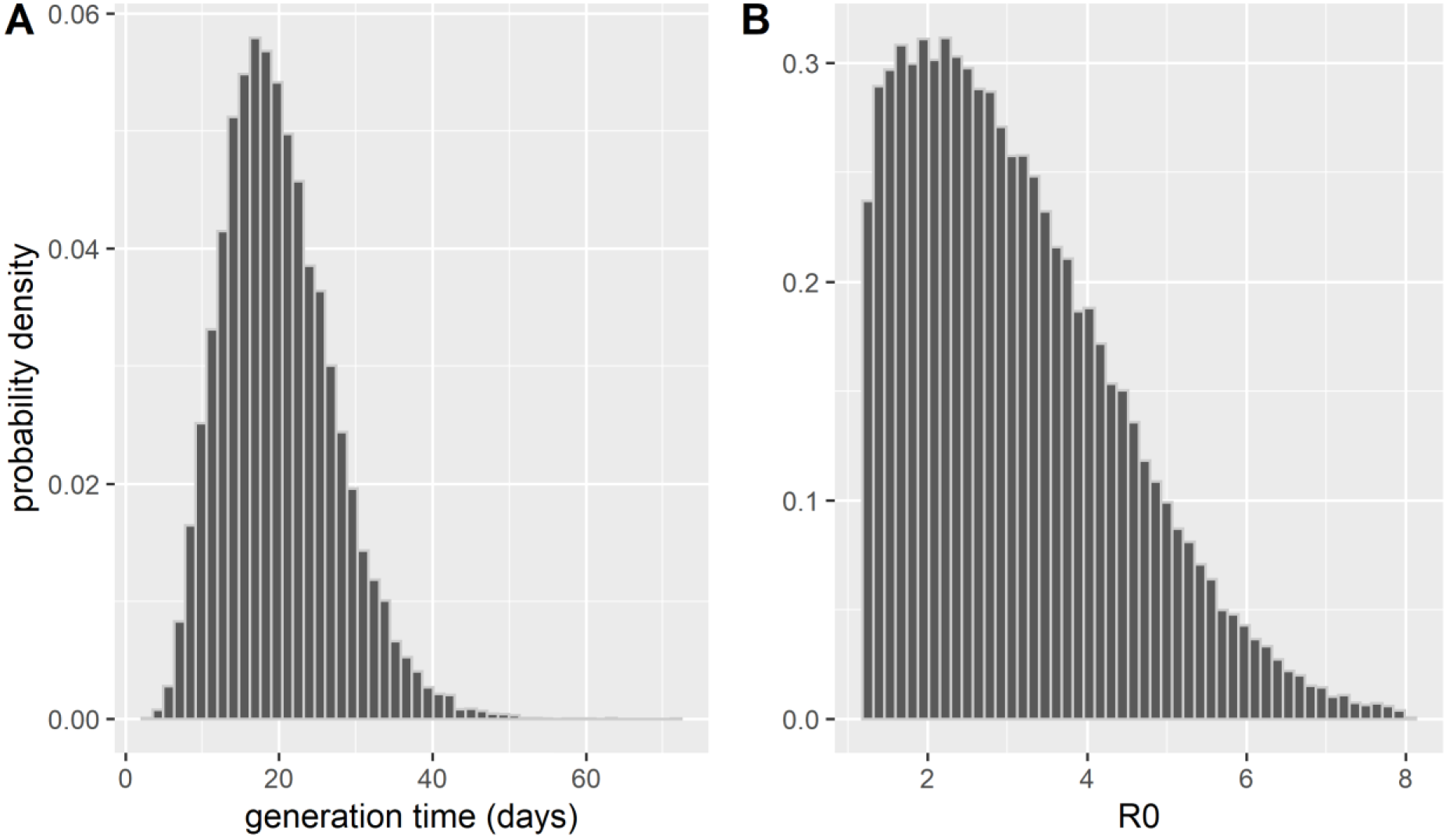
Distribution of the generation time and R0 values used to simulate epidemic data. Panel A shows the generation time distribution assumed in this analysis (gamma distribution with mean of 20 and standard deviation 7.4 days). Panel B shows the distribution of R_0_ values that was randomly drawn from for epidemic simulation (truncated normal distribution with mean of 2, standard deviation of 2, truncated between 1.2 and 8 days).

**Figure S2.**
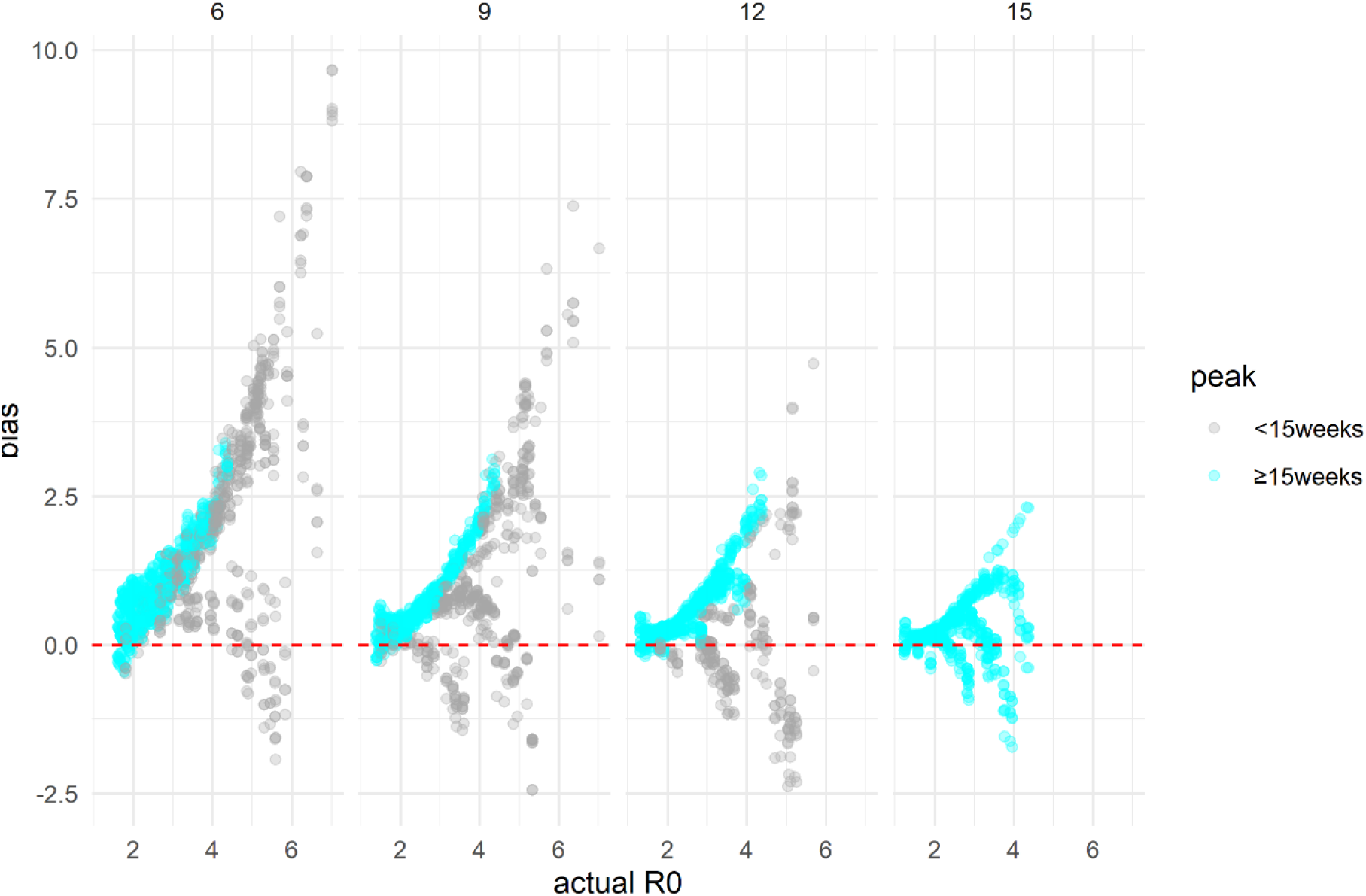
Distribution of bias in R0 estimates (estimated R0 – actual R0) when fitting to an increasing number of data points in the case time series of simulated datasets (N=245) without noise, pooled across methods. Columns represent the number of weeks fitted to in the case time series (6,9,12,15 weeks approximating to 2,3,4,5 generation times). Blue coloured points show the bias in estimates from simulations which peaked ≥15 weeks (N=146), i.e. highlighting values with a consistent distribution of true R_0_ values across the 4 time points assessed. Grey points show bias in estimates from simulations that peaked earlier than 15 weeks, therefore not represented across all time points assessed here. Red dashed lines indicate a bias value of 0, i.e. estimated R_0_ = actual R_0_.

**Figure S3.**
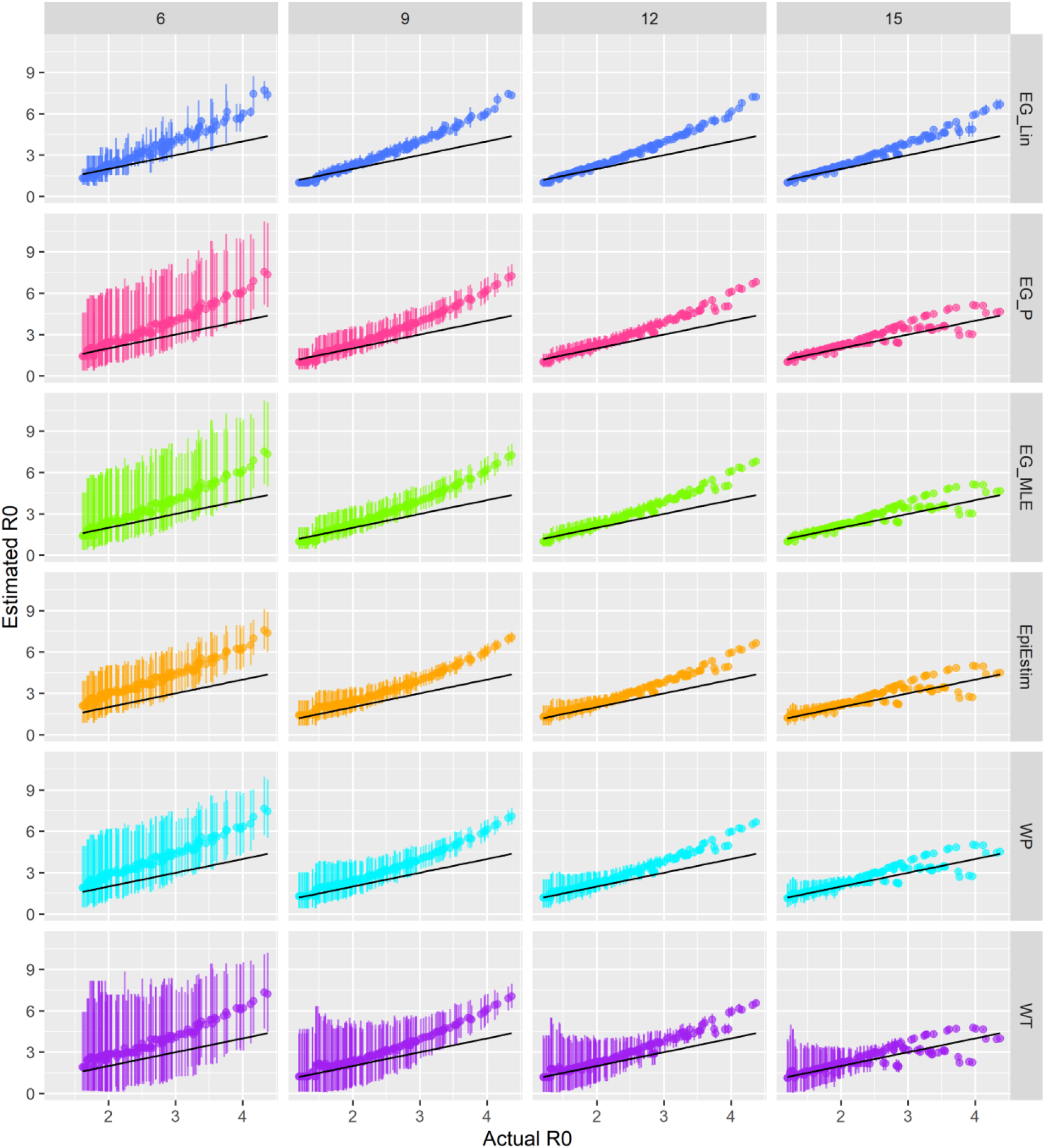
Comparison of estimated and actual R_0_ values from 146 simulated datasets that peaked ≥15 weeks, with no added noise. Columns show the number of weekly data each method was fitted to, and colours represent the method. Black lines represent the y=x line. Method abbreviations: Linear exponential growth rate method (EG_Lin); Poisson exponential growth rate method (EG_P); maximum likelihood exponential growth rate method (EG_MLE); White and Pagano method (WP); Wallinga and Teunis method (WT).

**Figure S4.**
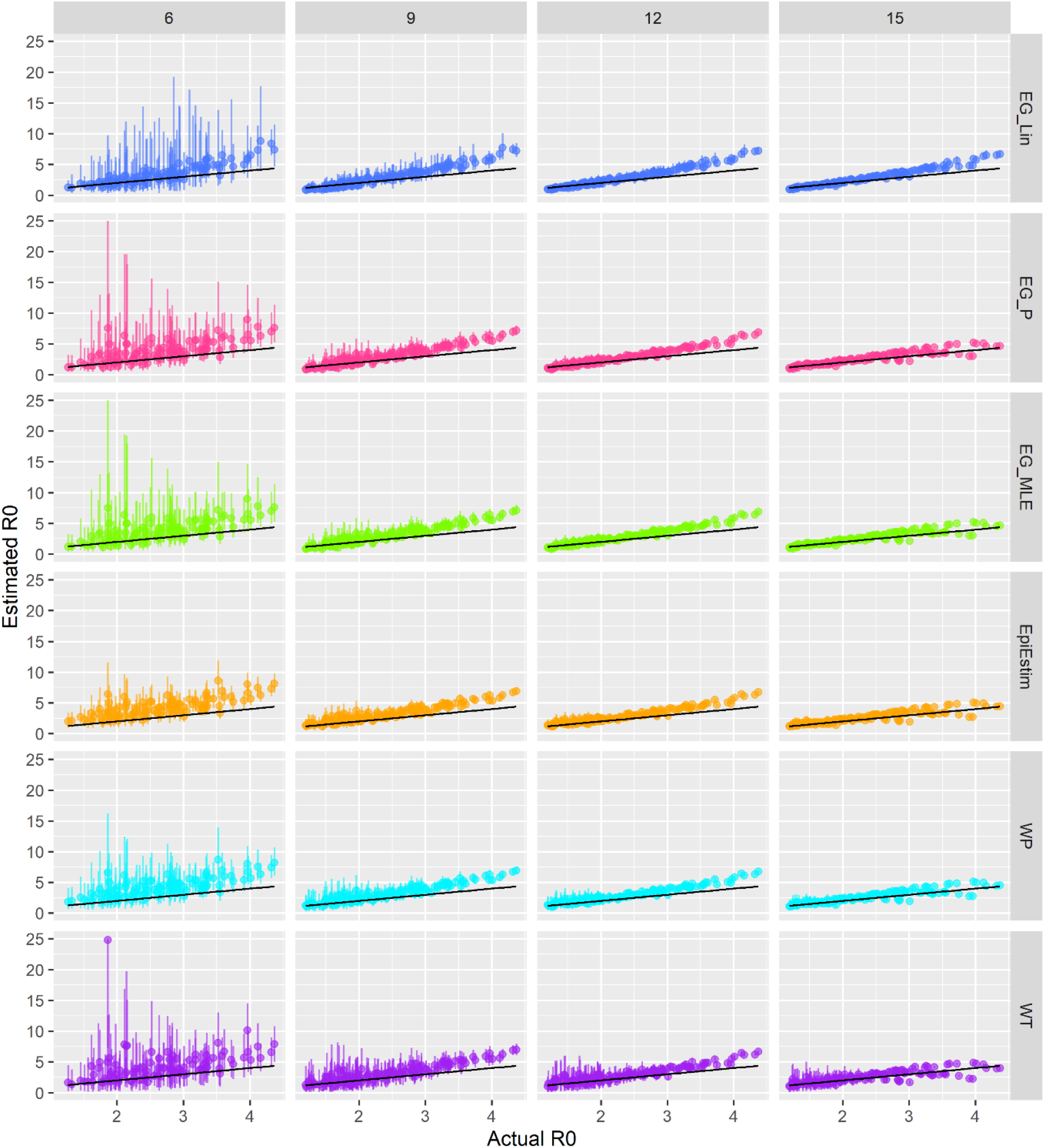
Comparison of estimated and actual R0 values from 148 simulated datasets that peaked ≥15 weeks, with Poisson random errors. Columns show the number of weekly data each method was fitted to, and colours represent the method. Black lines represent the y=x line. Method abbreviations: Linear exponential growth rate method (EG_Lin); Poisson exponential growth rate method (EG_P); maximum likelihood exponential growth rate method (EG_MLE); White and Pagano method (WP); Wallinga and Teunis method (WT).

**Figure S5.**
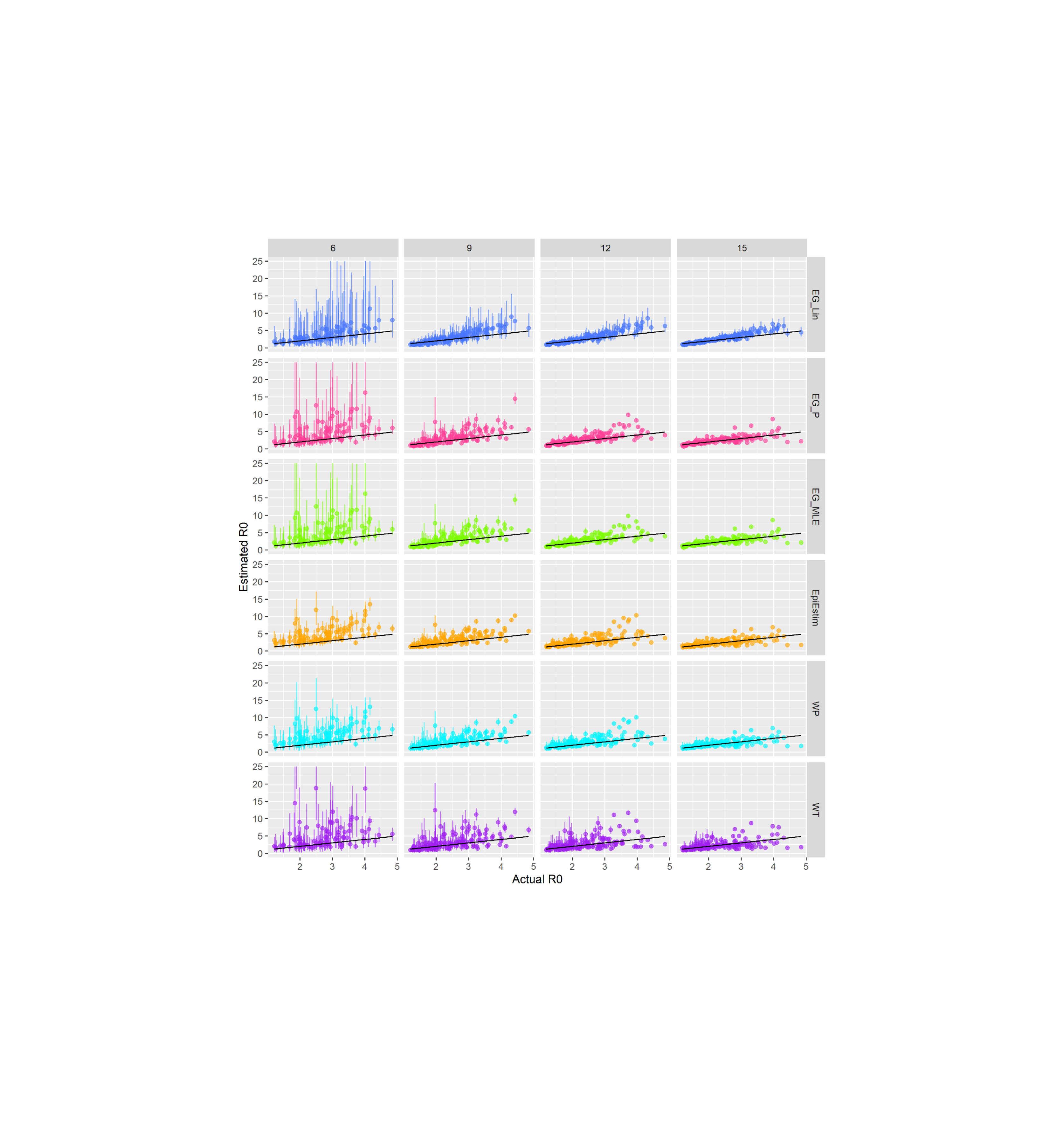
Comparison of estimated and actual R_0_ values from 152 simulated datasets that peaked ≥15 weeks, with negative binomial random errors. Columns show the number of weekly data each method was fitted to, and colours represent the method. Black lines represent the y=x line. Method abbreviations: Linear exponential growth rate method (EG_Lin); Poisson exponential growth rate method (EG_P); maximum likelihood exponential growth rate method (EG_MLE); White and Pagano method (WP); Wallinga and Teunis method (WT).

**Figure S6.**
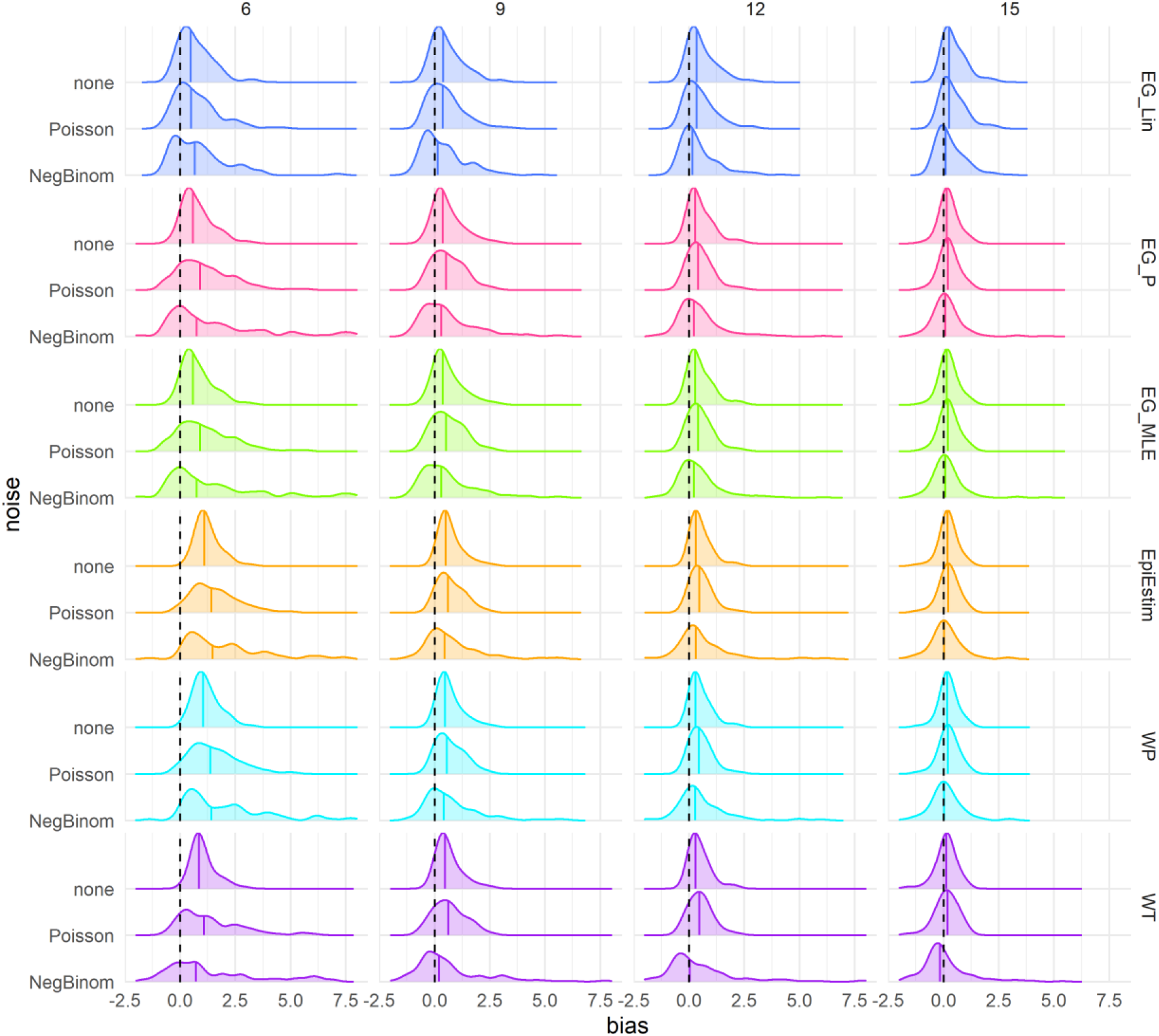
Density distributions of bias in R_0_ estimates (estimated R0 – actual R_0_) obtained when fitting to the case time series of simulated data, by method, time point (in approximate generations), and noise scenario, using only results from simulations that peaked ≥15 weeks (N=146). Columns represent the number of datapoints (weeks) each method was fitted to in the case time series (6,9,12,15 weeks). Black dashed lines highlight the ideal bias value of zero and coloured lines represent method-specific values of median bias. Method abbreviations: Linear exponential growth rate method (EG_Lin); Poisson exponential growth rate method (EG_P); maximum likelihood exponential growth rate method (EG_MLE); White and Pagano method (WP); Wallinga and Teunis (WT).

**Figure S7.**
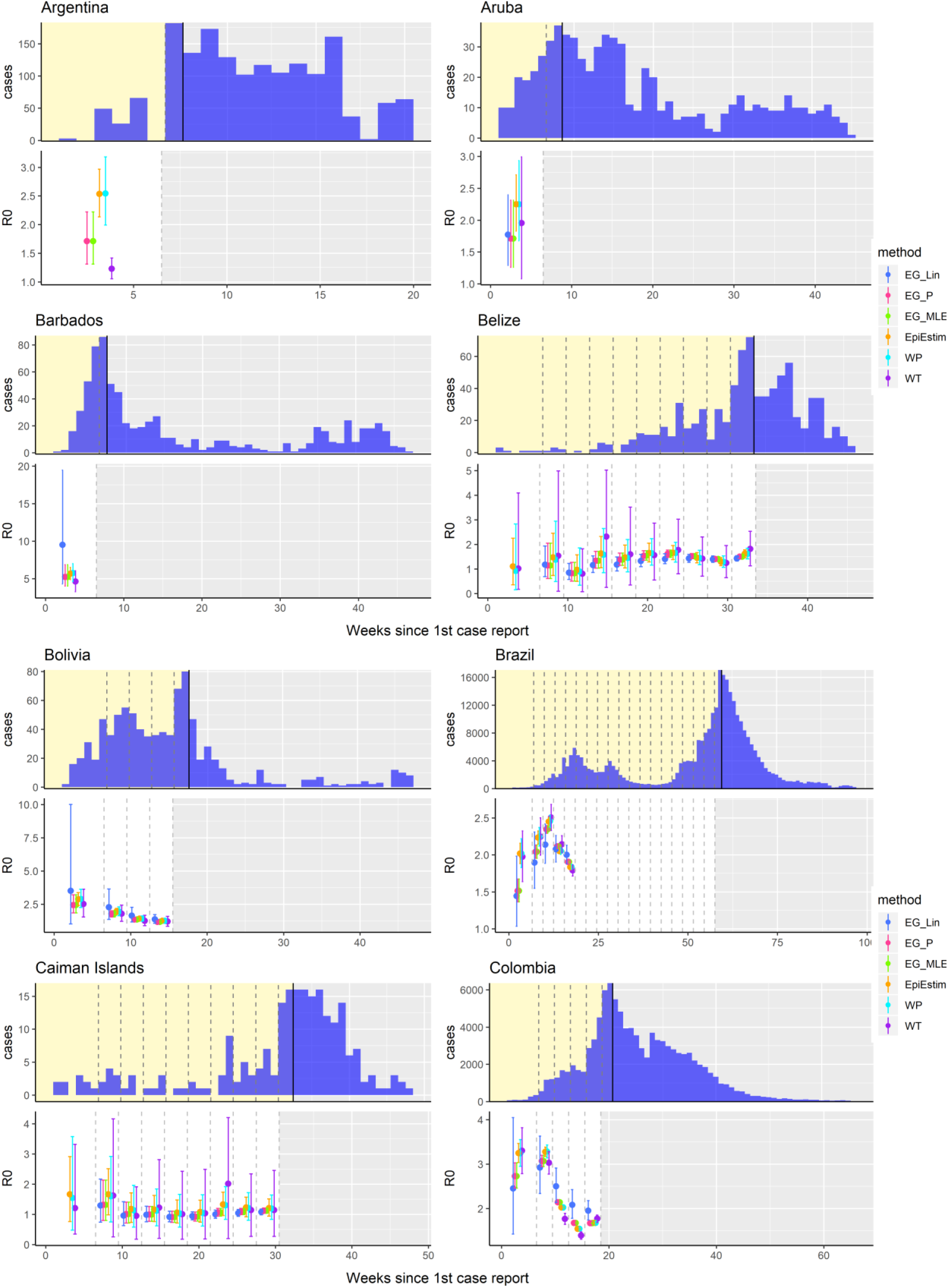
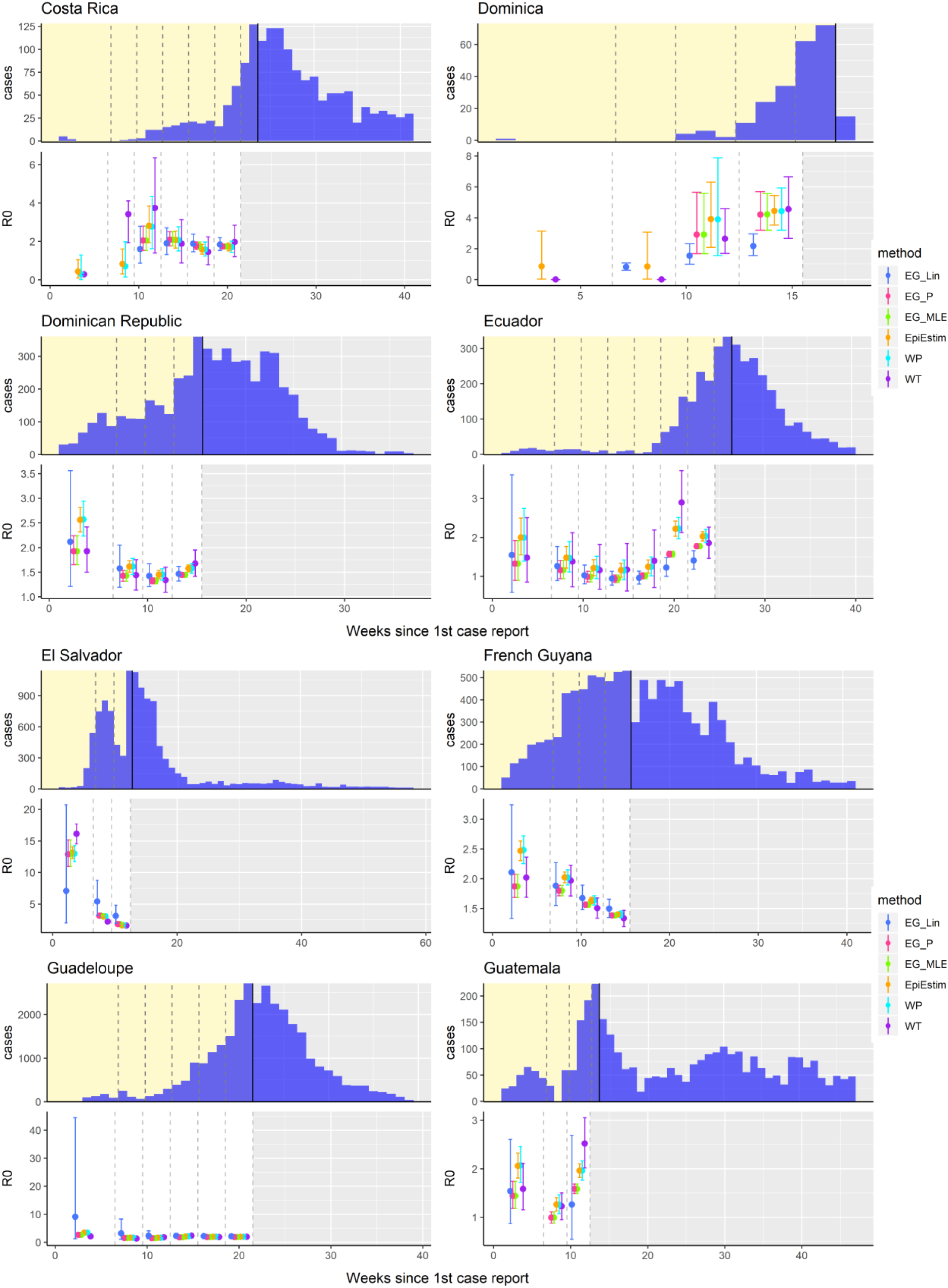
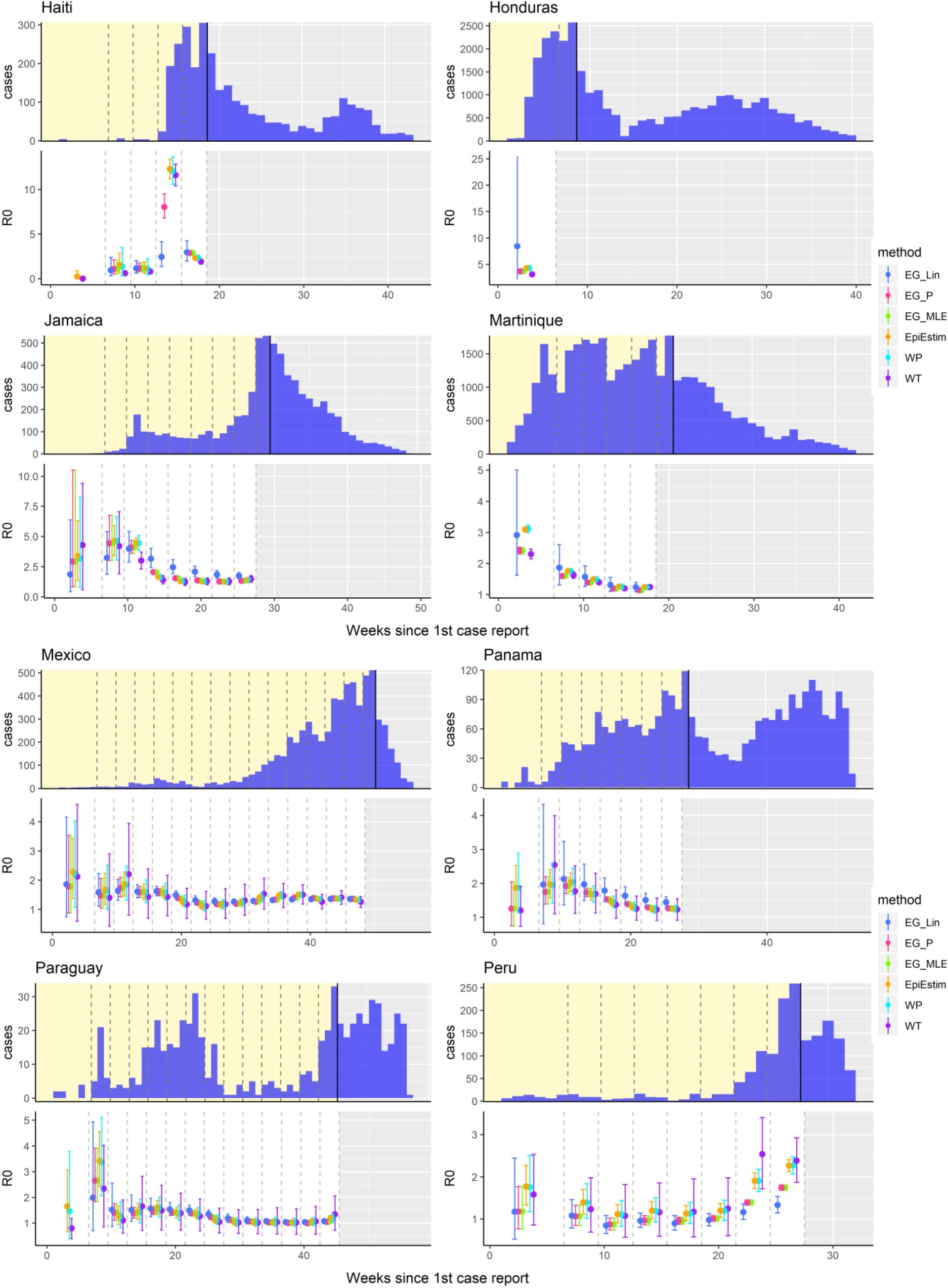
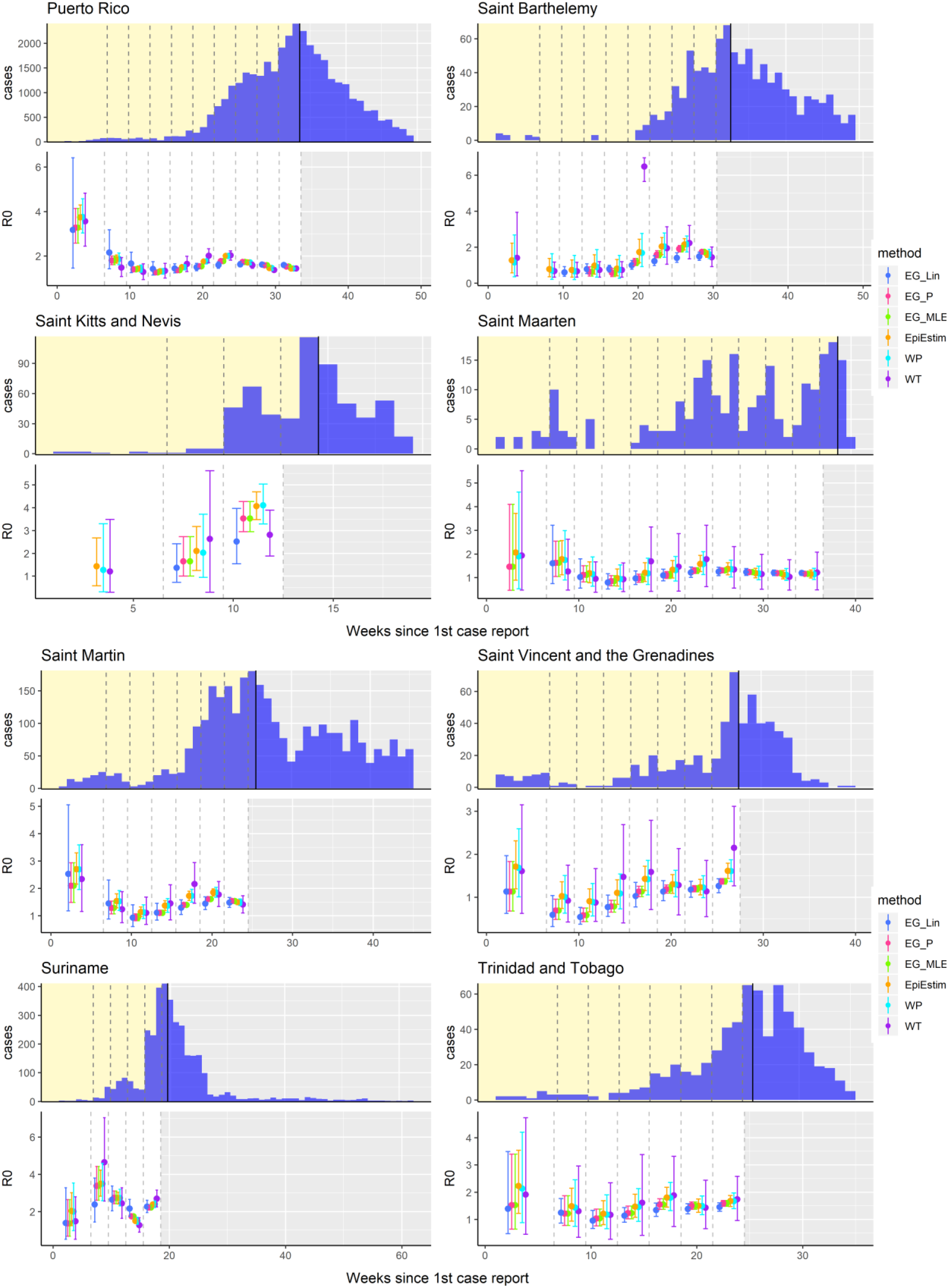
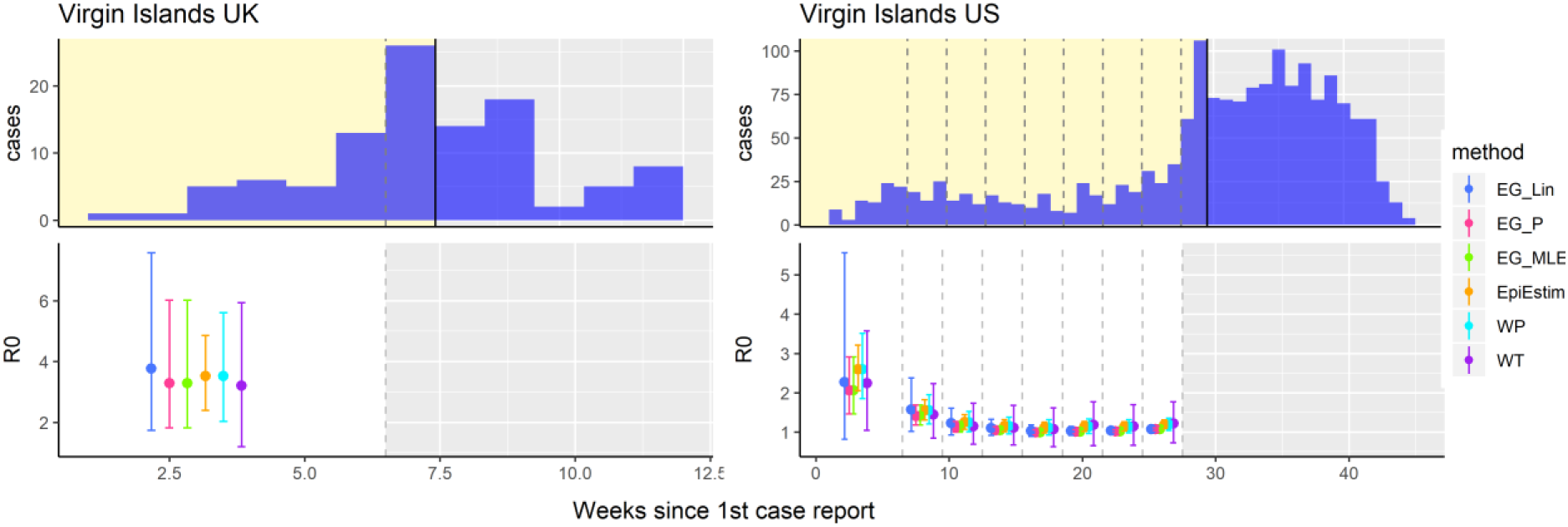
R_0_ estimates obtained from each of the six methods fitted at different stages of the epidemic growth phase from the 2015–2016 Zika epidemics in Latin America and the Caribbean. The top panel for each country shows the time series of reported Zika cases, with dashed lines showing the different stages at which each method was fitted to the data (first 6, 9, 12, etc., weeks) up to the peak of the epidemic, marked by the black line. The bottom panel for each country shows the mean and 95% confidence intervals of the R_0_ estimates produced with each method fitted to each time series. Method abbreviations: Linear exponential growth rate method (EG_Lin); Poisson exponential growth rate method (EG_P); maximum likelihood exponential growth rate method (EG_MLE); White and Pagano method (WP), Wallinga and Teunis (WT).

